# Prolonged Detection of SARS CoV2 RNA in Extracellular Vesicles in Nasal Swab RT-PCR Negative Patients

**DOI:** 10.1101/2021.12.16.21267912

**Authors:** P. Debishree Subudhi, Sheetalnath Rooge, Swati Thangariyal, Sivang Goswami, Reshu Agarwal, Ekta Gupta, Sukriti Baweja

**Affiliations:** Department of Molecular and Cellular Medicine, Institute of Liver and Biliary Sciences, Delhi, India; Department of Virology, Institute of Liver and Biliary Sciences, Delhi, India; Department of Transfusion Medicine, Institute of Liver and Biliary Sciences, Delhi, India

## Abstract

**Background:** There is a prolonged RT-PCR positivity seen in COVID-19 infected patients up to 2-3 months. We aim to investigate the presence of viral particles inside Extracellular vesicles (EV) and its role in underlying liver disease patients.

**Methods:** SARS CoV2 nasal RT-PCR positive n=78 {n=24(66.6%) chronic liver disease (CLD); n=52 (81.3%) non-liver disease} were studied. SARS CoV2 patients were also followed up for day (d) 7, 14 and 28. Extracellular vesicles were isolated using differential ultracentrifugation. SARS CoV2 RNA was measured using qRT-PCR by Altona Real Star kit.

**Results:** In baseline RT-PCR positive patients, SARS-CoV2 RNA inside the EV was present in 64/74 (82%) patients with comparable viral load between VTM and EV (mean 1CT – 0.033±0.005 vs. 1CT – 0.029±0.014, p=ns). On follow-up at day 7, of the 24 patients negative for COVID19, 10 (41%) had persistence of virus in the EV (1CT – 0.028±0.004) and on day 14, 14 of 40 (35%) negative RT-PCR had EVs with SARS CoV2 RNA (1CT – 0.028±0.06). The mean viral load decreased at day7 and day14 in nasal swab from baseline (p=0.001) but not in EV. SARS-CoV2 RNA otherwise undetectable in plasma, was found to be positive in EV in 12.5% of COVID19 positive patients. Interestingly, significantly prolonged and high viral load was found in EV at day 14 in CLD-COVID19 patients compared to COVID19 alone (p=0.002). The high cellular injury was seen in CLD-COVID19 infected patients with significant high levels of EV associated with endothelial cells and hepatocytes than COVID19 alone (p=0.004; 0.001).

**Conclusion:** Identification of SARS-CoV2 RNA in EV, in RT-PCR negative patients indicates persistence of infection for and likely recurrence of the infection. EV associated RNA may determine the clinical course of subjects with undetectable SARS-CoV2 virus and this may also have relevance in management of chronic liver disease patients.

## INTRODUCTION

A new coronavirus (CoV) identified as COVID-19 virus is the etiological agent responsible for the 2019-2021 viral pneumonia outbreak that commenced in Wuhan (1). The Covid-19 illness has a broad range of clinical manifestations, from asymptomatic or mild infection to a severe respiratory illness progressing to respiratory and multi-organ failure (2). Though the virus predominantly involves respiratory tract epithelium but its existence beyond respiratory tract is not yet explored. Reactivation/reinfection of the virus or prolonged detection of SARS-CoV2 could be due to simultaneous co-existence of the virus in extra-respiratory sites.

Despite many reports have characterized the clinical, epidemiological, laboratory, and radiological features, as well as treatment and clinical outcomes (3,4) of patients with COVID-19 pneumonia, the information of the SARS-CoV-2 reactivation remains a mystery. The curative and eradicative therapy for COVID-19 is not currently available. Urgent questions that need to be addressed promptly include whether patients with COVID-19 pneumonia are getting the reactivation or recurrence. The present detection tools have its own limitation and the risk factors predict SARS-CoV-2 reactivation or prolonged detection in patients is not been known. Therefore, there could be false negatives on occasion for oropharyngeal or nasopharyngeal swabs tests, affected by the site from which the sample was taken, the experience of the operator, and the actual quantity of virus (5, 6).

Recently, we have shown in case of hepatitis B that even in patients who were hepatitis B virus DNA negative, inside their extracellular vesicles, the HBV DNA was present with low amounts. We also found that extracellular vesicles associated with HBV DNA were able to infect and transmit the virus in naïve hepatocytes (7).

Presently, the reasons for COVID19 recurrence are largely unknown. Primarily due to its abrupt presentation not related to age, sex, or other physiological parameters (8). There is an urgent need to detect the virus in any associated form it is present from where the virus could may contribute in reactivation or recurrence so that complete cure of this infection can be achieved. Extracellular vesicles (EVs), including microvesicles (MVs), or exosomes have been shown to serve as vehicles for intercellular communication and transfer of genetic material in several viral infections (9). Therefore, it is reasonable to hypothesize that EVs may serve as reservoirs of SARS Cov2, transmit COVID-19 infection to naÏve cells and the EV associated SARS Cov2 RNA in future might also contribute in reactivation after the viral clearance.

Therefore, in the present study, we intend to identify the SARS-CoV2 RNA in associated form with extracellular vesicles in both nasal swab and plasma. In the present study, we took advantage of a widely established centrifugation based extracellular vesicle purification strategy and characterized the relationship between SARS CoV2 RNA and EVs with the follow up of patients, when the RNA is undetectable in nasal swab. And, whether the EV associated SARS CoV2 is able to transmit infection in naÏve cells in vitro.

## PATIENTS AND METHODS

### Patients

Patients infected with SARS-CoV2 were enrolled between December 2020-May 2021. COVID-19 patients (>-18 years) were selected based on a confirmed diagnosis on PCR assays (2 viral genes, envelope protein or E gene and RNA-dependent RNA polymerase or RdRP gene) of nasopharyngeal swab samples. COVID-19 infections were categorised as mild, moderate and severe. Patients with cirrhosis plus bacterial sepsis hospitalized in the intensive care unit (ICU) of ILBS were also included in the study. SARS CoV2 nasal and throat swab RT-PCR positive n=78 {n=24chronic liver disease (CLD); n=52 non-liver disease} n=5 RT PCR negative subjects (HC) were studied. SARS CoV2 patients were also followed up for day (d) 7, and 14. Nasal swab [collected in viral transport media (VTM)] and plasma samples were investigated at each time point. All patients were negative for other viral infections, including hepatitis D virus, HBV, HCV and HIV, and had no autoimmune liver diseases, sepsis or septic shock or patients on ventilator support. The clinical characteristics of the COVID19 patients with and without liver disease are summarized in Table.1. Institutional Ethics committee approved the study (F.37/ (1)/9/ILBS/DOA/2020/20217/513) and patients were enrolled after taking informed written consent. Biobank samples were collected as per the ILBS biobank guidelines, and samples from in-house hospitalized patients and hospital staff were obtained after proper informed consent from the patient or his/her relative following the national bioethical guidelines.

**Table.1.** Baseline characteristics of COVID19 patients

| Variable | COVID19 Alone | COVID19 +CLD | p-Value |
| --- | --- | --- | --- |
| Age (years) | 54 (34–65) | 24 (34–78) | n.s. |
| Male, n (%) | 46 (86.3) | 19 (85) | n.s. |
| Neutrophil to lymphocyte ratio | 8.15 (2.5–39.3) | 7 (4.4–18) | n.s. |
| Platelet count ($\times 10^9/L$) | 210 (57–449) | 71 (22–143) | <0.0001 |
| Bilirubin (mg/dl) | 0.77 (0.33–3.74) | 2.91 (0.63–29.8) | 0.009 |
| AST (U/L) | 54 (27–210) | 55 (23–273) | n.s. |
| ALT (U/L) | 68.5 (18–172) | 63 (21–101) | n.s. |
| Albumin (g/dl) | 3.29 (0.37) | 2.8 (0.57) | 0.001 |
| Serum sodium (mEq/L) | 131.5 (4.03) | 134 (7.24) | n.s. |
| INR | 1.05 (0.96–1.74) | 1.4 (0.98–2.54) | 0.0002 |
| Creatinine (mg/dl) | 0.9 (0.53–1.65) | 1.19 (0.4–3.4) | n.s. |
| ICU, n (%) | 11 (20) | 12 (50) | n.s. |
| ARDS, n (%) | 12 (54.5) | 9 (45) | n.s. |
| Liver disease severity n (%) | – |  |  |
| Child-Pugh A | 0 (0) | 2 (8.3) |  |
| Child-Pugh B | 0 (0) | 5 (20) |  |
| Child-Pugh C | 0 (0) | 18 (25) |  |
| MELD score | – | 22 (16–28) | -- |
| Comorbidities n (%) |  |  |  |
| Hypertension | 9 (16.6) | 4 (16.6) | n.s. |
| Diabetes mellitus | 11 (20.3) | 9 (37.5) | n.s. |
| Coronary artery disease | 5 (9) | 3 (12.5) | n.s. |
| Obesity | 2 (3.7) | 3 (12.5) | n.s. |

### Extracellular Vesicles Isolation Using Differential Ultracentrifugation

Circulating EVs were isolated from plasma and nasal swab [collected in viral transport media (VTM)] using differential ultracentrifugation. The collected pellet was then characterised on basis of size, morphology and origin of cells. Detailed protocol of sample collection, processing and isolation were followed according to the special guidelines by ISTH for extracellular vesicles to avoid any artifacts as described previously by our group (10).

### Characterization of COVID19 Associated Extracellular Vesicles using Transmission Electron Microscopy

To confirm the size and shape of the EVs, transmission electron microscopy was performed. 2-5µl of EVs sample was placed on the formvar carbon-coated, copper grids and incubated for 10 mins. After 10 minutes, excess sample was washed twice with 100µl of decarbonated water and dried at room temperature for 2 mins. Grids were then examined in Talos F200C transmission electron microscopy (Thermo Scientific) and capture using software/camera/device.

### Quantification of Extracellular Vesicles using Nanoparticle Tracking Assay (NTA)

EVs were quantification and sizes were determined using Nanosight 3000. EVs from plasma and nasal swab were diluted in 1:500 ratio in endotoxin free PBS. 60s video was recorded and analysed using NTA 3.1 Build 3.1.46 version, camera level 15 and detection threshold at 3. Temperature was monitored during recording. The mean size (nm) concertation (particles/ml) was calculated and plotted as particle size versus number of particles per mL.

### Cell of Origin of Circulating Extracellular Vesicles Using Multicolor Flow Cytometer

For the detection of EVs by flow cytometer, we used spherotech latex beads (IL, USA), a mixture of microbeads of four different sizes (0.22 μm, 0.44 μm, 0.88 μm, and 1.0 μm), to confirm the size of the EVs. Multiparameter flow cytometry was performed according to a standard protocol, and the data was acquired using a FACS Verse flow cytometer and analyzed using FlowJo software (Treestar Inc., Ashland, OR, USA). Epithelial cells (Epcam+ CK19+ CDh1+), endothelial cells (CD31+CD45-), and hepatocytes (ASGPRII+) surface markers and Annexin V (early apoptosis) was seen, as described in detail previously (10) were used as the markers of EVs to confirm the cell of origin.

### SARS CoV2 RNA Quantification

SARS CoV2 RNA was extracted from EVs and isolated and RT PCR was performed for E gene and RNA dependent RNA polymerase (RdRP) using one step Altona Real Star kit according to the manufacturer’s instructions. To remove any viruses derived that might be adsorbed on the cell surface, isolated EVs were treated with RNAse H (Sigma Aldrich, St Louis, MO, USA) before Viral RNA isolation.

### Co-infection of Vero Cells by Infected EVs from COVID19 patients in Cell Culture

Vero cells were cultured in DMEM medium (GIBCO/BRL, Grand Island, NY, USA), supplemented with 100 U/ml penicillin, 100 mg/ml streptomycin and 10% fetal bovine serum. The EVs isolated from CHB patients were co-infected with vero cell line (in ratio 1:10) and SARS CoV2 RNA was quantified at 12h, 24h and 72h after infection and MTT assay was performed to investigate the cytotoxicity.

### Statistical Analysis

All the data were analysed using GraphPad Prism 7 (GraphPad Software, Inc. La Jolla, CA, USA) and SPSS. When appropriate, the univariate statistical techniques applied were Student’s t-test/ Mann–Whitney-U test and one-way analysis of variance to determine statistically significant differences among the control and experimental groups. Fischer’s test was used to determine odds ratio. Significance was defined as p>0.05.

## RESULTS

### Baseline Clinical Characteristics of COVID19 Patients

A total of 78 COVID19 RT-PCR confirmed subjects were investigated, among them 52 (66.6%) patients were non-liver disease and 24 subjects (30.7%) were chronic liver disease. All the patients were followed up from baseline to day (d) 7, and 14. There was no significant difference among the groups with respect to age and sex. Among laboratory variables; neutrophil to lymphocyte ratio (NLR), aspartate aminotransferase (AST), alanine aminotransferase and sodium levels were not significantly different between COVID-19 and COVID-19 plus cirrhosis groups. Platelet count and albumin levels were lower while bilirubin and international normalized ratio (INR) were higher in the COVID-19 plus cirrhosis group. Platelet counts may be low in COVID-19 with cirrhosis due to effect of both liver disease and COVID-19 itself. As far as comorbidities were concerned, diabetes was the most common in both groups (20% in COVID-19 alone and 37.5% in COVID-19 plus CLD groups).

### Extracellular Vesicles Size and Morphological Characterization from COVID19 Patients

After isolation of EV from nasal swab (VTM) and plasma, were characterized using NTA assay and we found majority of EV were in range of 80-400nm in size and concentration was determined was between 2 X10 – 9 × 10 with no differences among two groups of COVID19 patients (Fig.1A). The morphology was determined using transmission electron microscopy which clearly shows the membranous structures in the size ranges between 75-380nm/ per field as visible at 73000x, 120,000x with the zeta potential mean of 27.1mV ±2.84. (Fig.1B and C).

**Figure.1.**
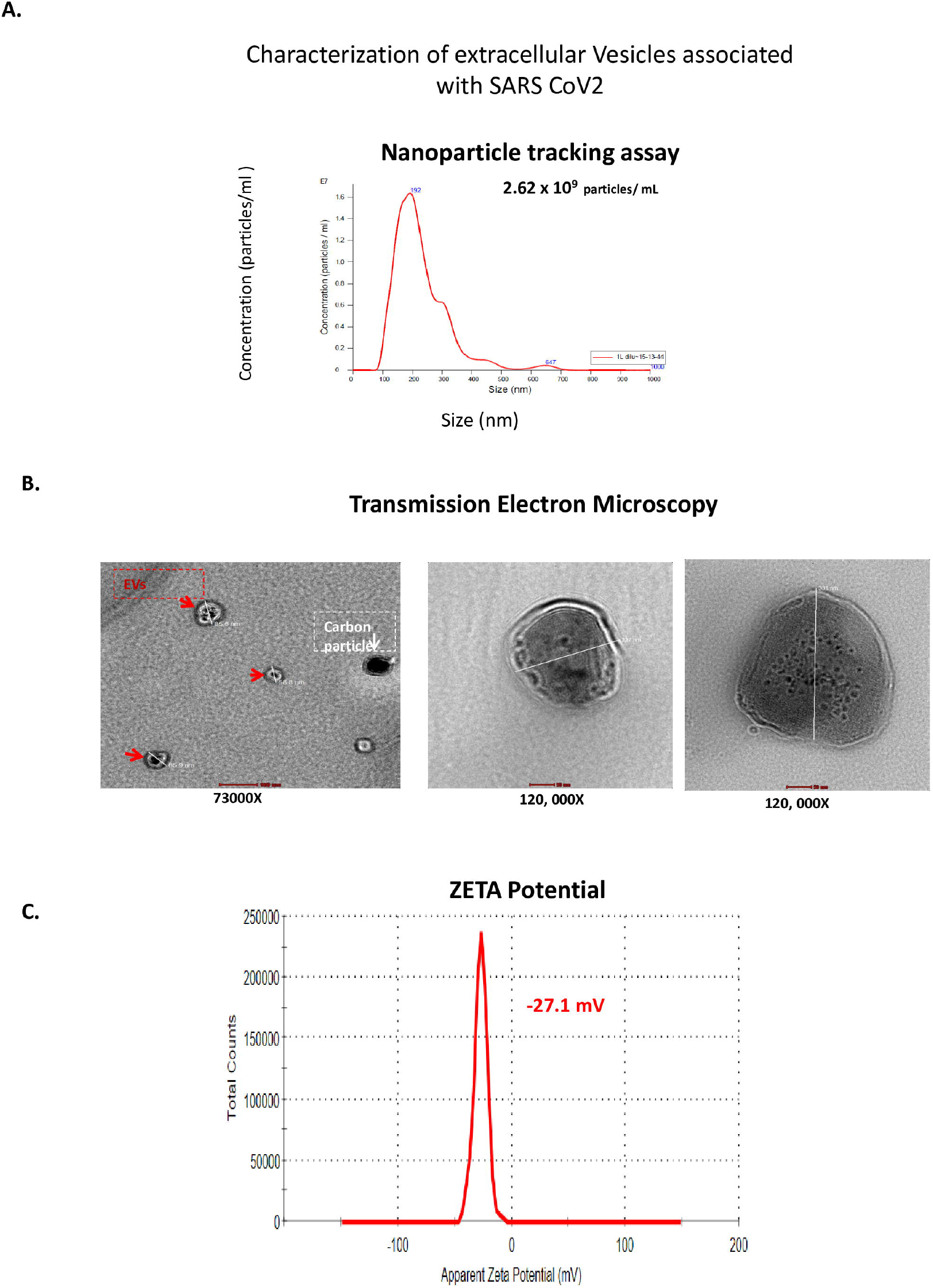

### Extracellular Vesicles Derived from COVID19 Patients Contained SARS-CoV2 RNA

The schematic representation of work flow is represented in Fig.2A. At the baseline (at the time of presentation with symptoms), nasal swab in VTM and EV isolated from VTM were analysed (Figure.2A). At baseline RT-PCR positive patients, SARS-CoV2 RNA inside the EV was present in 64/74 (82%) patients with comparable viral load between VTM and EV (mean 1CT – 0.033±0.005 vs. 1CT – 0.029±0.014, p=ns) (Figure.2B). The mean viral load decreased at day7 and day14 in nasal swab from baseline (p=0.001; 0.001 respectively), whereas no viral load differences were seen in EV (Figure.2B).On follow-up at day 7, of the 24 patients negative for COVID19, 10 (41%) had persistence of virus in the EV (1CT – 0.028±0.004) and on day 14, 14 of 40 (35%) negative RT-PCR had EVs with SARS CoV2 RNA (1CT – 0.028±0.06) (Figure.2C). SARS-CoV2 RNA otherwise undetectable in plasma. It was found to be positive inside EV in 12.5% of COVID19 positive patients and their EV minus plasma were negative for RNA (Fig.2D).

**Figure.2.**
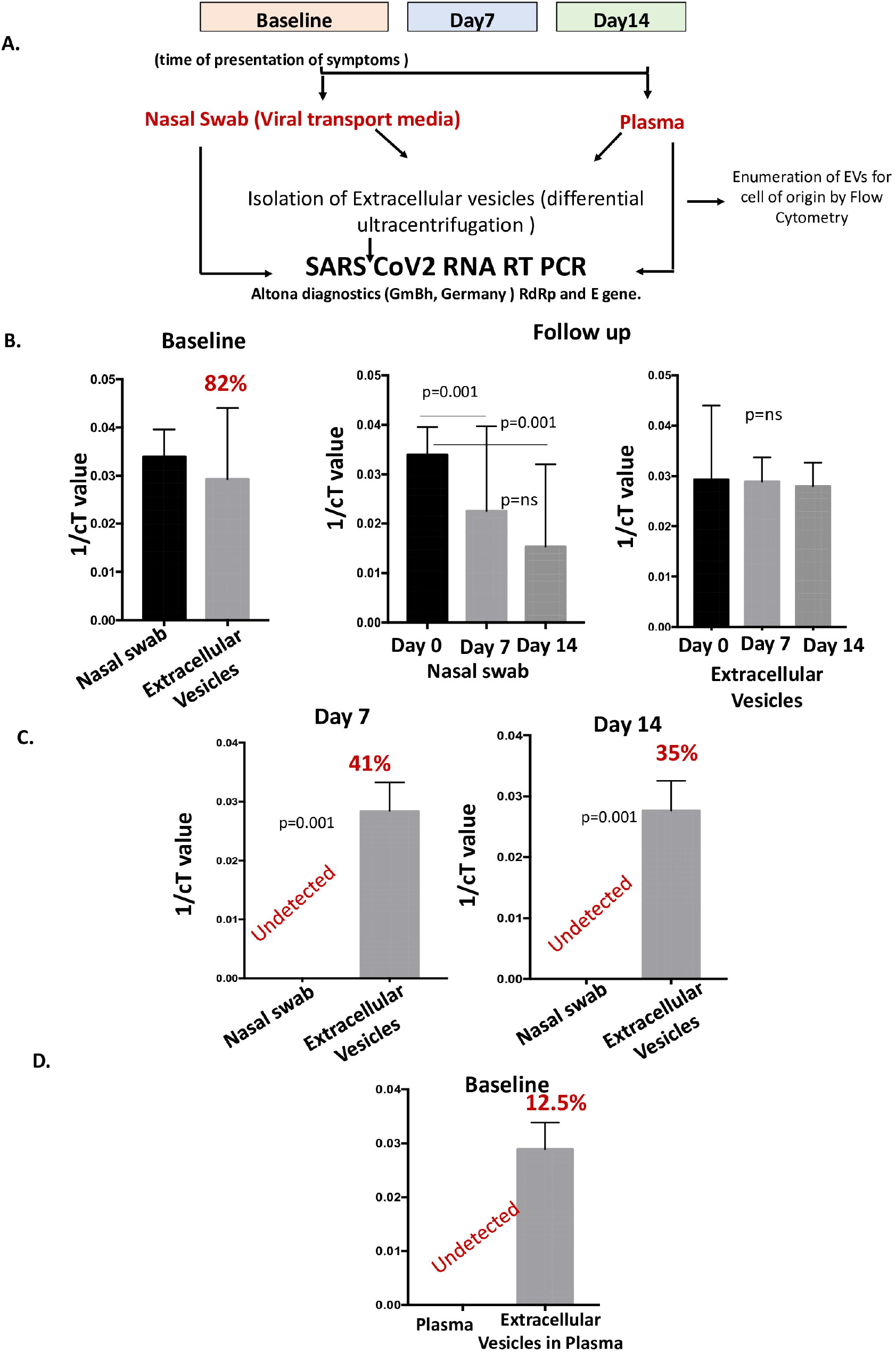

### Extracellular Vesicle Associated SARS CoV2 RNA in COVID19 with Chronic Liver Disease Patients

Interestingly, upon comparison among COVID19 infected patients with or without chronic liver disease, we found no significant differences in CT values in nasal swab VTM RNA, whereas, we found significantly prolonged and high viral load was found in EV at day 14 in CLD-COVID19 patients compared to COVID19 alone (p=0.002) (Fig.3)

**Figure.3.**
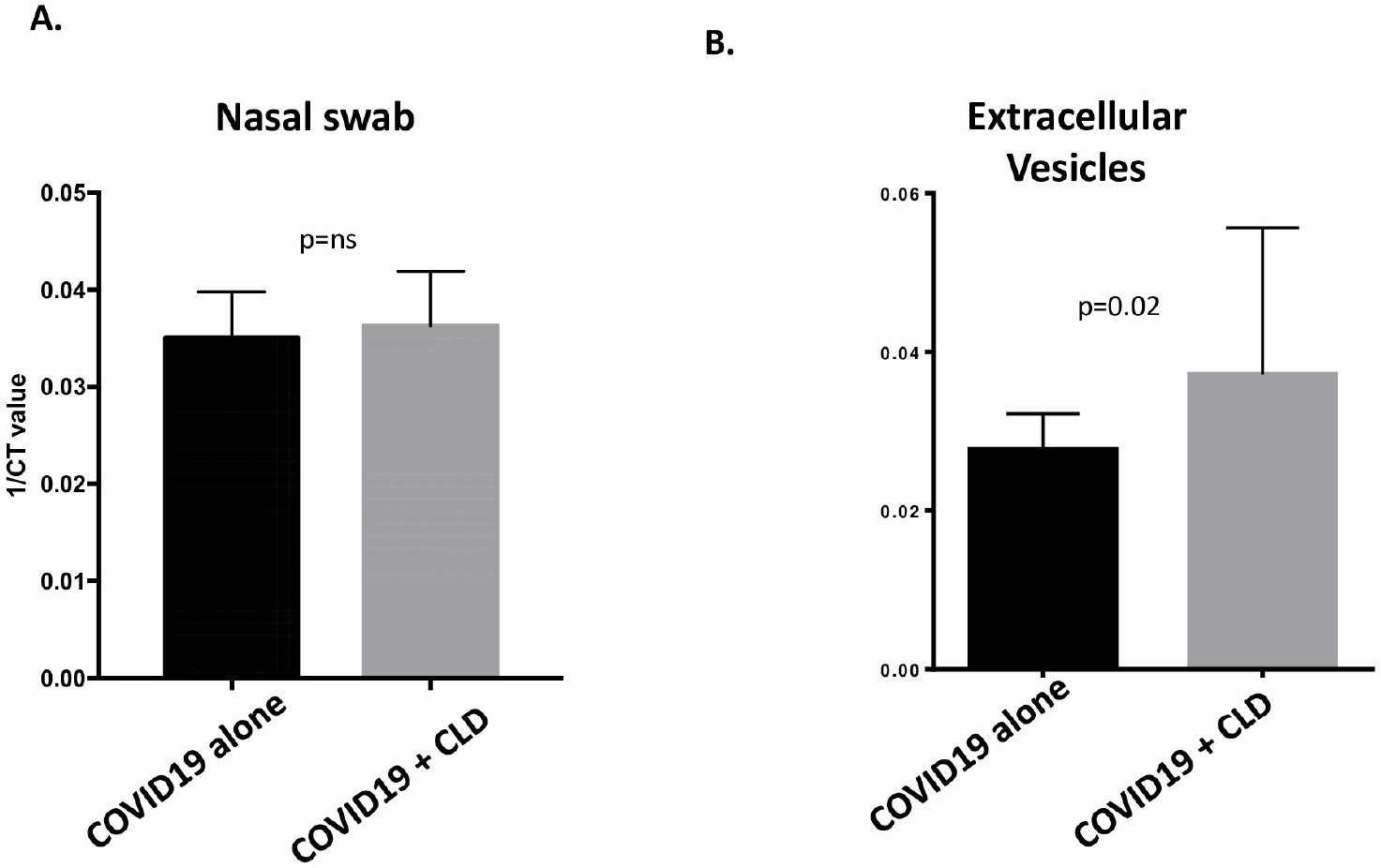

### Cellular Origin of Extracellular Vesicles in COVID19 patients

Next, in order to determine the cellular origin of EVs and cell injury in COVID19 patients, which we analysed EV and confirmed using epithelial cells (Epcam+ CK19+ CDh1+), endothelial cells using flow cytometry in all the groups. By using different latex size beads and Annexin V^+^, we were able to detect EVs Fig.4A is the representative dot blot of flow cytometry with gating at different sizes and counting beads used to analyse the origin of EVs. In COVID19 patients EV associated with epithelial, endothelial cells and hepatocytes, were high levels than healthy controls (p=0.001; 0.001., 0.001) (Fig.4B,C,D). Whereas, the high cellular injury was seen in CLD-COVID19 infected patients with significant high levels of EV associated with endothelial cells and hepatocytes than COVID19 alone (p=0.004; 0.001) (Fig.4C and D).

**Figure.4.**
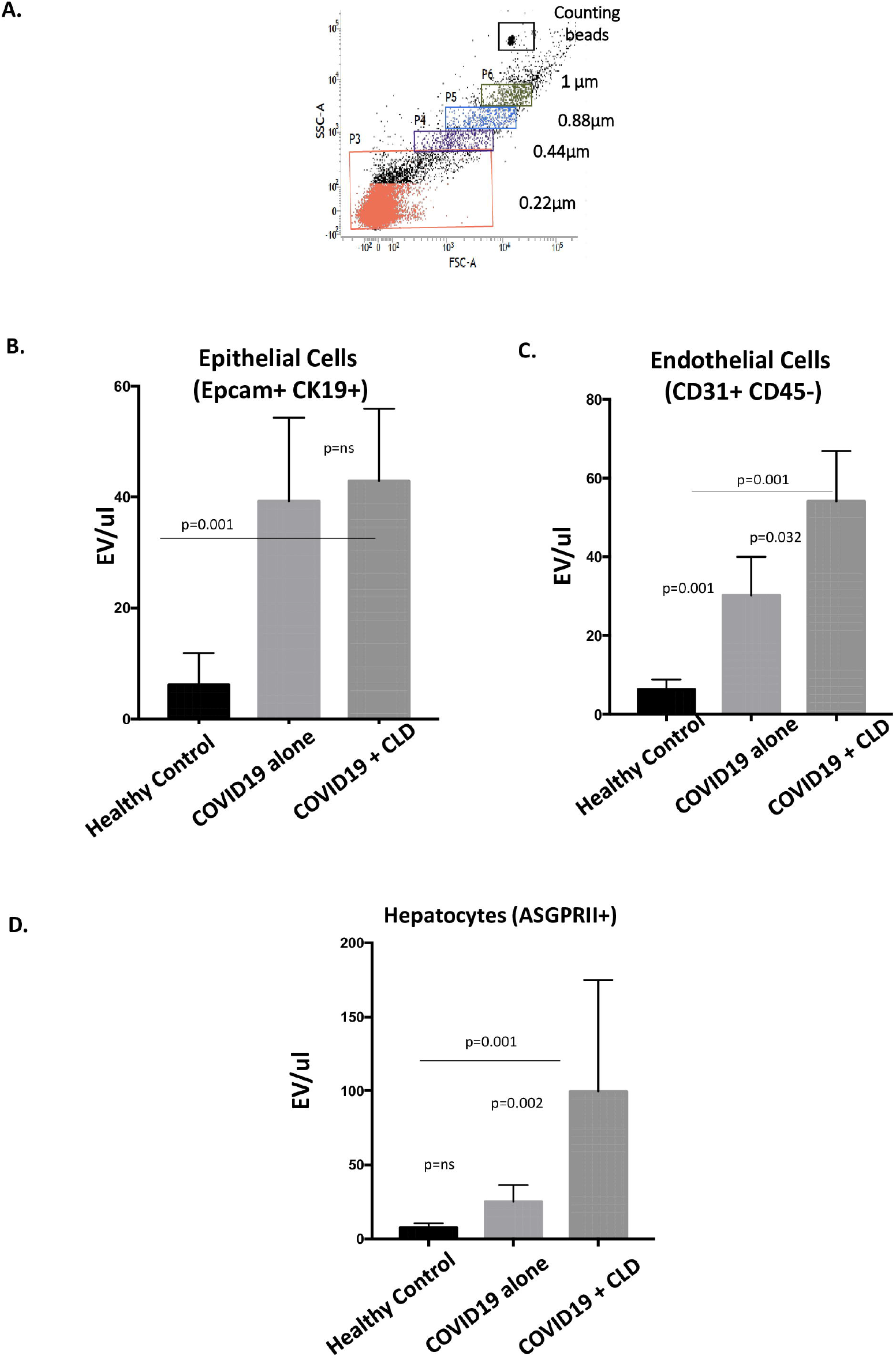

### Extracellular Vesicles Associated SARS-CoV2 RNA is Transmissible and Infectious

To understand that EVs from COVID19 patients can infect naïve cells, we infected Vero cells EVs from both the groups of COVID19 and determined the SARS-CoV2RNA at 0, 24, 48 and 72 hours in Vero cells. We found that EVs were able to transmit SARS CoV2 RNA into naïve cells 24h post-infection as suggested by SARS CoV2 RNA RT-PCR for E gene and RdRp gene. Further, SARS-CoV2 RNA levels in Vero cells increased with time as determined by CT Values (E-gene at 24h 20.1 ±5.6and 23.6±6.8 at 72h) indicating SARS-CoV2 RNA replication post-transmission from infected EVs (Fig.5B). Infected EV may induce apoptosis in immortalized cells, the effect of infected EV on Vero cells was assessed using an MTT assay. The treatment had no significant effect on the viability of Vero cells even after 24, 48 and 72h (Fig.5C).

**Figure.5.**
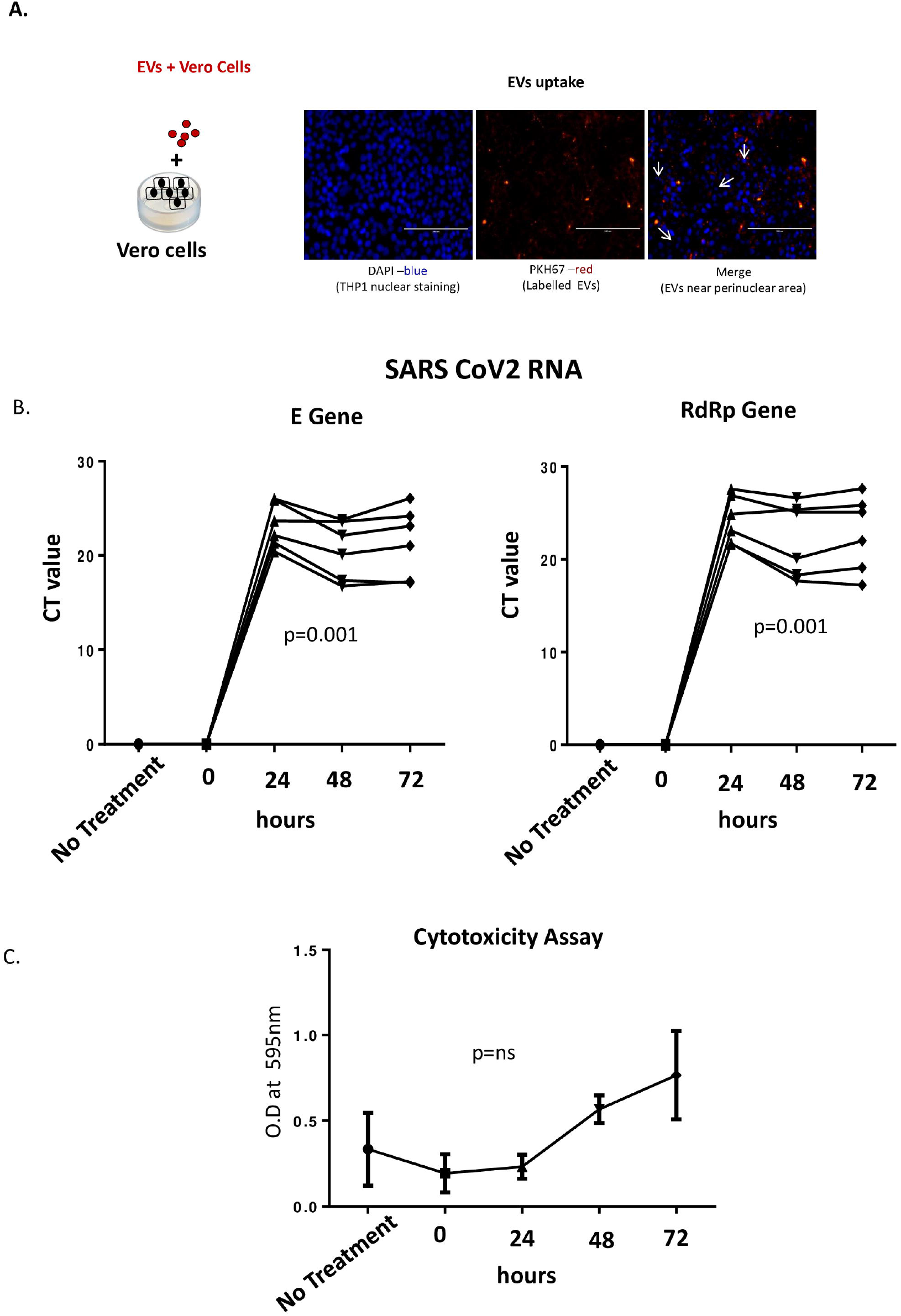

## DISCUSSION

The novelty of the study is that for the first time we identified SARS-CoV2 RNA in EV, in RT-PCR negative patients which indicates persistence of infection for and likely recurrence of the infection. Also, we found that infected EV are capable to transmit and infect the naïve Vero cells. It is suggestive of another route of transmission as EV harbour SARS CoV2 RNA. EV associated RNA may determine the ongoing inflammation and clinical course of subjects with undetectable SARS-CoV2 virus and this may also have relevance in management of chronic liver disease patients.

SARS-CoV-2 (Severe Acute Respiratory Syndrome Coronavirus 2; previously known as 2019-nCoV) cause of present pandemic since 2019. Presently, it remains to be determined the origins and possible intermediate animal vectors of SARS-CoV-2, as well as the mechanism that this virus spread among humans. Despite many reports have characterized the clinical, epidemiological, laboratory, and radiological features, as well as treatment and clinical outcomes (11,12) of patients with COVID-19 pneumonia, the information of the SARS-CoV-2 reactivation remains a mystery. The curative and eradicative therapy for COVID-19 is not currently available. Urgent questions that need to be addressed promptly include whether patients with COVID-19 pneumonia are getting the reactivation or recurrence. The present detection tools have its own limitation and the risk factors predict SARS-CoV-2 reactivation in patients is not been known. In a recent report it has been confirmed that in a significantly proportion of COVID-19 patients, SARS-CoV-2 reactivation developed after discharging from hospital (9%)(13). Also, reported the clinical characteristics of these patients with SARS-CoV-2 reactivation which were similar to those of non-reactivated patients with COVID-19 infection (14). In our findings, presence of SARS CoV2 in associated form with extracellular vesicles reveals for the first time the hidden form of RNA. Notably, based on few reports there is currently an evidence to suggest that a proportion of recovered COVID-19 patients again got COVID-19 positivity (15). We also found that even in RT-PCR negative patients, SARS-COV2 RNA was detectable even after 14 days inside the EVs.

The results of the SARS-CoV-2 RNA tests, in such cases are fluctuant. May be because, no research has yet accurately established the contagious period of COVID-19. Our study will be first of its kind, to identify the new route of transmission or infectivity. Besides patients and asymptomatic carriers, those in convalescence may also be infectious. SARS-CoV-2 RNA from respiratory tract specimens may be persistent or recurrently positive during the course of this disease (16,17).

Furthermore, Angiotensin-converting enzyme-2 (ACE-2), identified as the cell entry receptor of SARS-CoV-2, was highly expressed in the lungs rather than in the upper respiratory tract (18). The result of the SARS-CoV-2 RNA test likely depends on the viral load of the specimen. Therefore, there could be false negatives on occasion for oropharyngeal or nasopharyngeal swabs tests, affected by the site from which the sample was taken, the experience of the operator, and the actual quantity of virus. The Bronchoalveolar lavage fluid (BALF) specimen test is considered more accurate but with a higher exposure risk. In addition to the above specimens, SARS-CoV-2 RNA can be detected in a patient’s sputum, blood, or stool swab by RT-PCR assay (19). Running multiple tests and collecting different specimens would be more effective approaches to maximize sensitivity. Therefore, we propose to investigate the SARS-CoV-2 RNA in the extracellular vesicles and which will decipher the alternative form of infection and transmission of COVID-19 and it may pave way for newer and better prognostic tool to detect the COVID-19. Also, EVs are well known to be the miniatures of cells and every cell type releases them during the disease condition.

EVs are present in all the body fluids and in circulation in abundance. Hence, we found even in plasma, where it SARS-CoV-2 RNA was undetectable, it was found associated with EVs. Recently, many assays are developed using quantitative RT–PCR (qRT–PCR) approaches for detection of the virus. However, the typical turnaround time for screening and diagnosing patients with suspected SARS-CoV-2 has been >24⍰h, given the need to ship samples overnight to reference laboratories. Although serology tests are rapid and require minimal equipment, their utility may be limited for diagnosis of acute SARS-CoV-2 infection, because it can take several days to weeks following symptom onset for a patient to mount a detectable antibody response. EV associated SARS CoV2 RNA detection may solve the above problems. Therefore, there is an urgent public health need for rapid and more precise diagnostic tests for SARS-CoV-2 infection.

Identification of SARS-CoV2 RNA in EV, in RT-PCR negative patients indicates persistence of infection for and likely recurrence of the infection is suggestive of another route of transmission as EV harbour SARS CoV2 RNA. Therefore, it is worthwhile to propose that in case of COVID-19 where we are unable to understand the mechanism of recurrence or false negativity, it could be possible that we may find the virus associated with EVs and we can easily detect in circulation and could find out the reasons for major problem which we foresee of reactivation and its associated mortality.

## Data Availability

All data produced in the present study are available upon reasonable request to the authors

